# Hierarchical neurobiological changes in dorsolateral prefrontal cortex and subgenual anterior cingulate cortex connectivity induced by theta-burst stimulation

**DOI:** 10.1101/2025.05.23.25328212

**Authors:** Armin Toghi, Ghazaleh Ghaffaripour Jahromi, Shayan Zarei, Babak Aliyari, Hamed Moqtaderi, Ali Motie Nasrabadi

## Abstract

Recent studies show that left dorsolateral prefrontal cortex (DLPFC) stimulation induces acute and persistent changes in the subgenual anterior cingulate cortex (sgACC), and the degree of sgACC modulation predicts treatment response in patients. To understand how theta-burst stimulation (TBS) over the left DLPFC influences sgACC microcircuitry, we applied spectral dynamic causal modeling (DCM) with a conductance-based canonical microcircuit model (CMM-NMDA) to resting-state EEG data acquired at 2, 15, and 30 minutes following continuous TBS (cTBS), intermittent TBS (iTBS), and Sham stimulation. Using a multilevel parametric empirical Bayes framework, we observed that cTBS induce wider network effect 2 min after stimulation while iTBS made more lasting changes up to 30 min following stimulation. Unlike sham, both TBS protocols led to a transient reduction in NMDA-dependent effective connectivity from right DLPFC to left sgACC. Notably, TBS caused sustained inhibition within left sgACC microcircuits, characterized by increased self-inhibition of spiny-satellite cells. Together, these findings demonstrate that spectral DCM can resolve the dynamic, hierarchical neuromodulatory effects of TBS in healthy brains and highlight a potential mechanism underlying the antidepressant action of left DLPFC TBS.

## Introduction

Repetitive transcranial magnetic stimulation (rTMS) has emerged as a powerful tool in both neuroscientific research and clinical applications^1^. This technique uses focal electromagnetic fields to induce neuronal action potentials in the brain^1^. A more recent advancement in rTMS is theta burst stimulation (TBS), which delivers magnetic pulses in a pattern that mimics natural brain oscillations ^2^. Specifically, it consists of rapid gamma-frequency bursts (50 Hz) nested within slower theta-frequency oscillations (5 Hz) ^2–4^. Previous works showed that intermittent TBS (iTBS) enhance cortical excitability, while continuous TBS (cTBS) have an inhibitory effect on cortical activity ^3,5^.

The effectiveness of TBS lies in its ability to induce lasting neural modulation beyond the stimulation period^2^. While its neuroplastic effects have been most extensively studied in the human primary motor cortex (M1), the underlying mechanisms by which TBS modify activity in nonmotor regions remain unclear^4,6,7^. This is particularly true for the left dorsolateral prefrontal cortex (DLPFC), a crucial region for therapeutic interventions. Despite the precise mechanisms of TBS still being under investigation, the technique has received FDA approval for MDD treatment ^8^. However, a significant proportion of MDD patients do not respond to TBS, potentially due to poor understanding of TBS neural effects and optimal stimulation parameters^9^.

Recent studies consistently highlight the relationship between the remote regulatory effects of left DLPFC rTMS on the subgenual anterior cingulate cortex (sgACC) and its role in the treatment improvement ^1,10–12^. Aberrant connectivity between the DLPFC and sgACC has been implicated in the pathophysiology of depression across various neuroimaging modalities^13^, and strong evidence suggests that this connectivity is altered following rTMS ^1,9–11,14–16^. Notably, effective stimulation sites tend to have higher anticorrelation^10–12,17^, and denser polysynaptic structural connection with the sgACC^18^. However, several challenges remain for understanding the left DLPFC TBS effect on sgACC activity and connectivity.

First, using functional magnetic resonance imaging (fMRI) to track prefrontal TBS effects yields mixed findings^19^, commonly attributed to high intra- and inter-subject variability^3,19^. Moreover, applying concurrent TMS–fMRI to a full TBS protocol to track its network effect is challenging due to insufficient temporal signal-to-noise ratio (tSNR) in subcortical regions^20^. Second, electrophysiological studies showed that TBS does not produce a uniform inhibitory or excitatory response^6^, highly state-dependent^21^, induces variable and complex changes in spectral power^4^, and a triggers widespread network connectivity effects^9^. Third, TMS creates a complex chain of causation, spanning synaptic changes to broader network and behavioral outcomes^22^, that current methodological techniques cannot easily capture.

Instead, computational models with biologically plausible assumptions offer a bridge between synaptic changes and electrophysiological observations^23–25^. In particular, dynamic causal modeling (DCM) using conductance-based neural mass models of cortical microcircuitry has become a popular approach for examining treatment-associated changes in extrinsic interregional coupling, intrinsic connectivity, and cellular properties in each neural population^24,26^. This approach enables tracking hierarchical neurobiological effects of neuromodulation at multiple scales—macroscale (extrinsic interregional connections), mesoscale (intrinsic microcircuit interactions), and microscale (cellular properties) ^23,24^ **(Figure 1)**.

**Figure 1.**
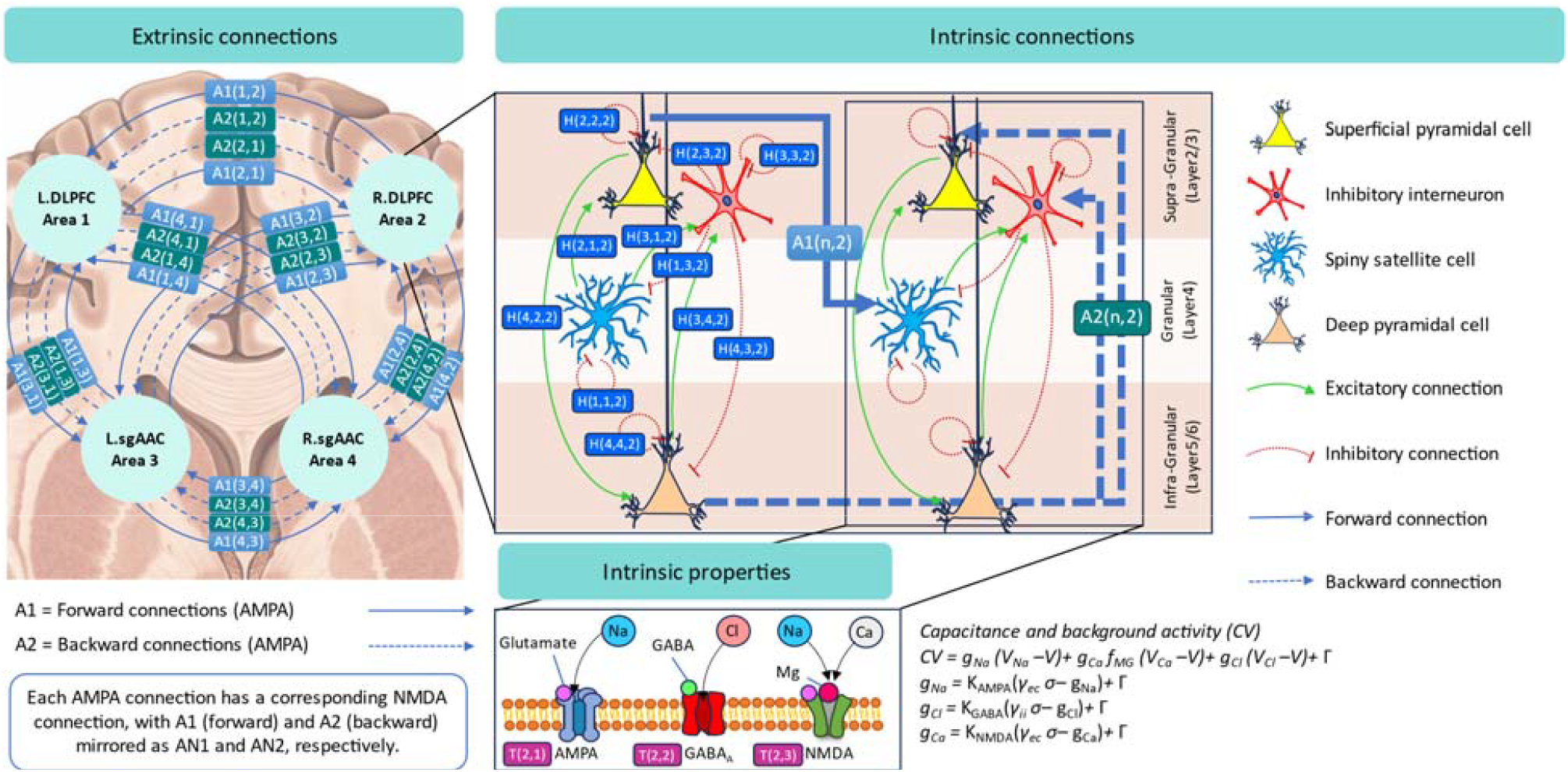
Hierarchical schematic of the CMM_NMDA neural model across three nested scales. (i) macroscale extrinsic forward (A1) and backward (A2) AMPA-mediated connection (same for NMDA) between four sources; (ii) mesoscale intrinsic laminar circuitry within each source—superficial pyramidal, spiny satellite, inhibitory interneurons and deep pyramidal cells across supra-granular, granular and infragranular layers 2 with excitatory (green) and inhibitory (red) interactions; and (iii) microscale conductance-based receptor and ion-channel dynamics (AMPA, NMDA, GABA □) that are defined at the source level—rather than per individual neuron—to capture aggregate postsynaptic currents.

DCM inverts a generative neural mass model alongside an observation model to infer intrinsic and extrinsic connectivity parameters that best explain the empirical data^27^. Spectral DCM adopted for resting state data to estimate how endogenous neuronal fluctuations produce the observed CSD^27^, and has demonstrated strong reliability in resting-state electrophysiology^28^. In this study, we used spectral DCM under a conductance-based canonical microcircuit model (CMM_NMDA) to investigate TBS-induced effective connectivity changes between the bilateral DLPFC and sgACC. Resting-state EEG data were recorded in a placebo-controlled, single-blinded, crossover design involving 24 healthy adult participants, each completing three sessions (iTBS, cTBS, sham). EEG recordings were obtained at baseline and at 2, 15, and 30 minutes post-stimulation **(Figure 2)**.

**Figure 2.**
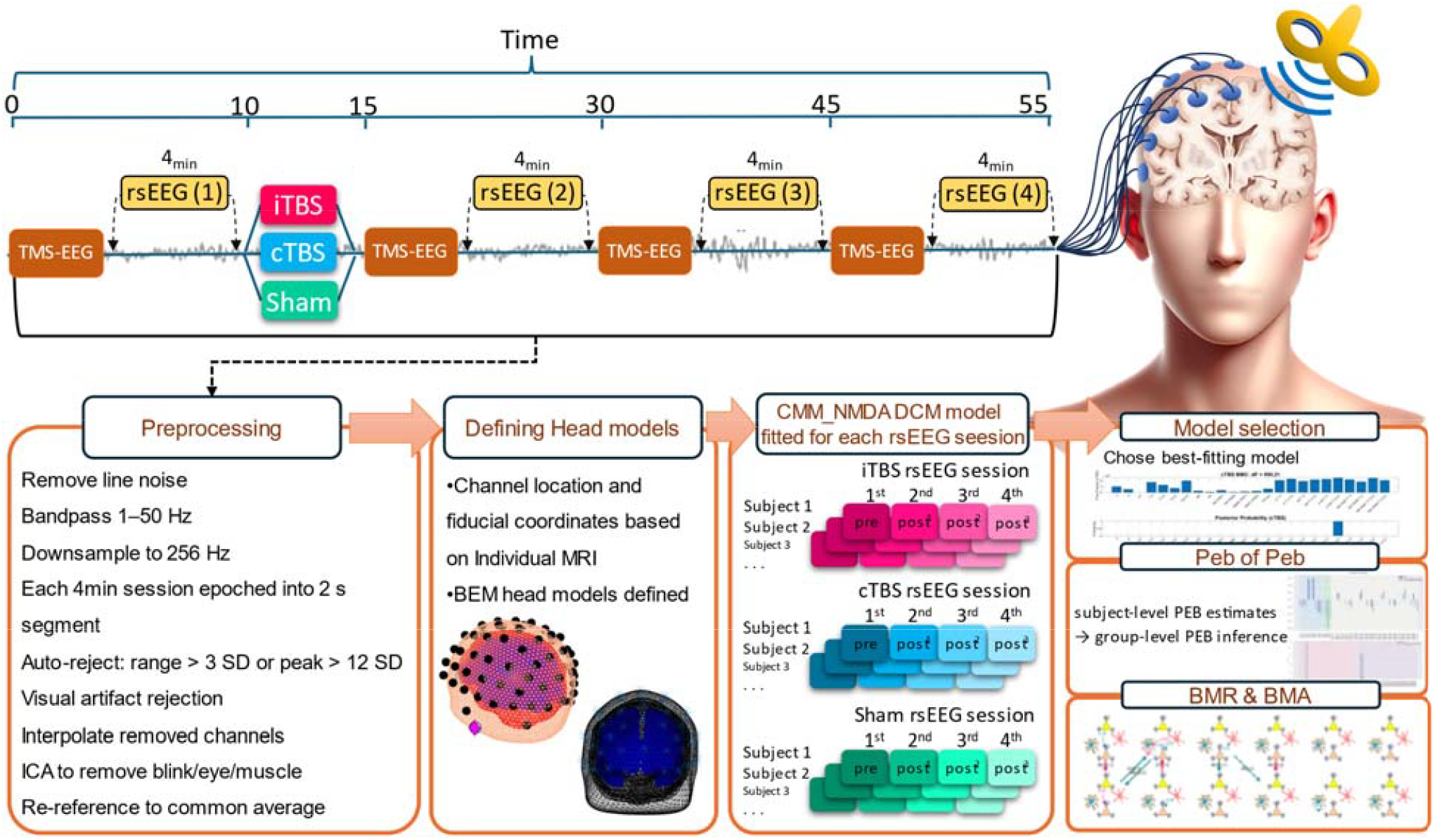
Overview of experimental design and analysis pipeline. Each participant completed three single-blinded TMS-EEG sessions (iTBS, cTBS, sham; randomized crossover, ≥1 week apart). In each session, 4 min of eyes-open resting-state EEG was recorded before TBS and after each of four single-pulse TMS blocks (pre, post^1^, post^2^, post^3^). EEG data were preprocessed and cleaned. Subject-specific T1-weighted MRIs were used to build three-layer BEM head models and cortical meshes. Source-level cross-spectral densities were computed and inverted in SPM25 using the conductance-based CMM_NMDA microcircuit model for each time block. Hierarchical parametric empirical Bayes used for group level analysis and Bayesian model reduction and averaging were used as an efficient form of model selection.

After model inversion for each session, we applied a two-level hierarchical Bayesian approach^24,29^ to assess the network effects of iTBS, cTBS, and sham at 2 minutes and to model subsequent dynamic changes up to 30 minutes. This method is a multilevel Bayesian estimation of a general linear model, previously used in longitudinal DCM studies^24,29^, which have two level optimizations over posterior densities. At the first level, empirical priors from individual level optimize posterior densities over parameters at the session level. Then, group-level means serve as empirical priors that constrain parameter estimates at the individual level (**Figure 3**). Crucially, this multilevel framework is particularly well-suited to account for the substantial intra- and inter-subject variability inherent to TMS interventions, and finesses the local minima problem inherent in the inversion of neural mass models (for the detailed description of this approach compared to classical statistics such as ANOVA see **Method** section).

**Figure 3.**
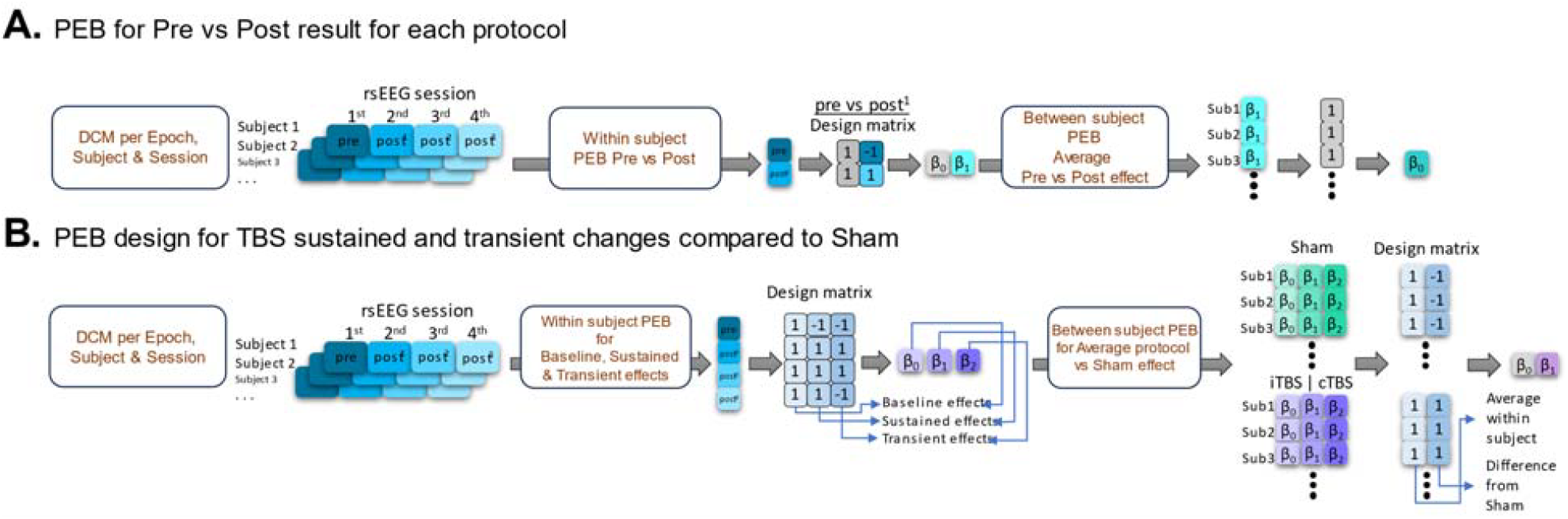
Group-level analysis using PEB of PEB. (A) Within-subject PEB uses a 2×2 matrix [1 −1; 1 1] to estimate each subject’s Pre→Post^1^ effect (β^1^), followed by a second-level PEB with a single-regressor [1] to compute the group mean differences (β^0^). (B) Within-subject PEB fits a 4×3 matrix across the four EEG blocks (pre, post^1^, post^2^, post^3^) with regressors for baseline [1 1 1 1], sustained change [−1 1 1 1] and transient change [−1 1 1 −1] to yield β (baseline), β^1^ (sustained) and β^2^ (transient). A second-level PEB then contrasts TBS (iTBS or cTBS) against sham using [1 −1; 1 1] to isolate TBS-specific modulations.

## Results

In group-level analyses we only considered connections with posterior probability > 0.95, reflecting strong evidence. We focus here on the difference connectivity matrices, where positive values denote increased connectivity and negative values denote decreased connectivity. Mean connectivity results are provided in the **Supplementary Material**.

### TBS effect 2 min after stimulation

Using the PEB-of-PEB framework (**Figure 3A**), we compared Pre versus Post1 (2 min) connectivity changes for iTBS, cTBS, and sham. Sham produced no significant extrinsic changes, as expected. In contrast, cTBS induced the largest extrinsic connectivity modulations: a significant increase in NMDA-mediated forward connectivity from left DLPFC to left sgACC. For iTBS, we observed the opposite pattern—a decrease in forward NMDA connectivity from left DLPFC to left sgACC and reduced backward AMPA coupling from left DLPFC to right sgACC. Across extrinsic connections, cTBS also yielded bidirectional reductions between left sgACC and right DLPFC, whereas iTBS did not significantly alter this pathway.

Intrinsic changes in the stimulation target further differentiated protocols: cTBS decreased SP → DP excitatory connectivity in left DLPFC, indicating local hypoactivation, while iTBS reduced DP→ II, reflecting local disinhibition. To quantify subject-level consistency, we extracted the sign of these two intrinsic parameters from each subject’s first-level PEB. Eighty-two percent of participants showed the inhibitory effect of cTBS, and 77% exhibited the disinhibitory effect of iTBS on left DLPFC microcircuit.

Additional intrinsic effects were found in sgACC and right DLPFC for cTBS only: left sgACC displayed increased SP→ DP, and right sgACC showed reduced II→DP inhibition. The sole significant change after sham was reduced SS self-inhibition in left sgACC. These immediate connectivity alterations highlight protocol-specific modulation of prefrontal–subgenual circuits and underscore the utility of DCM in resolving neuromodulatory effects (**Figure 4**).

**Figure 4.**
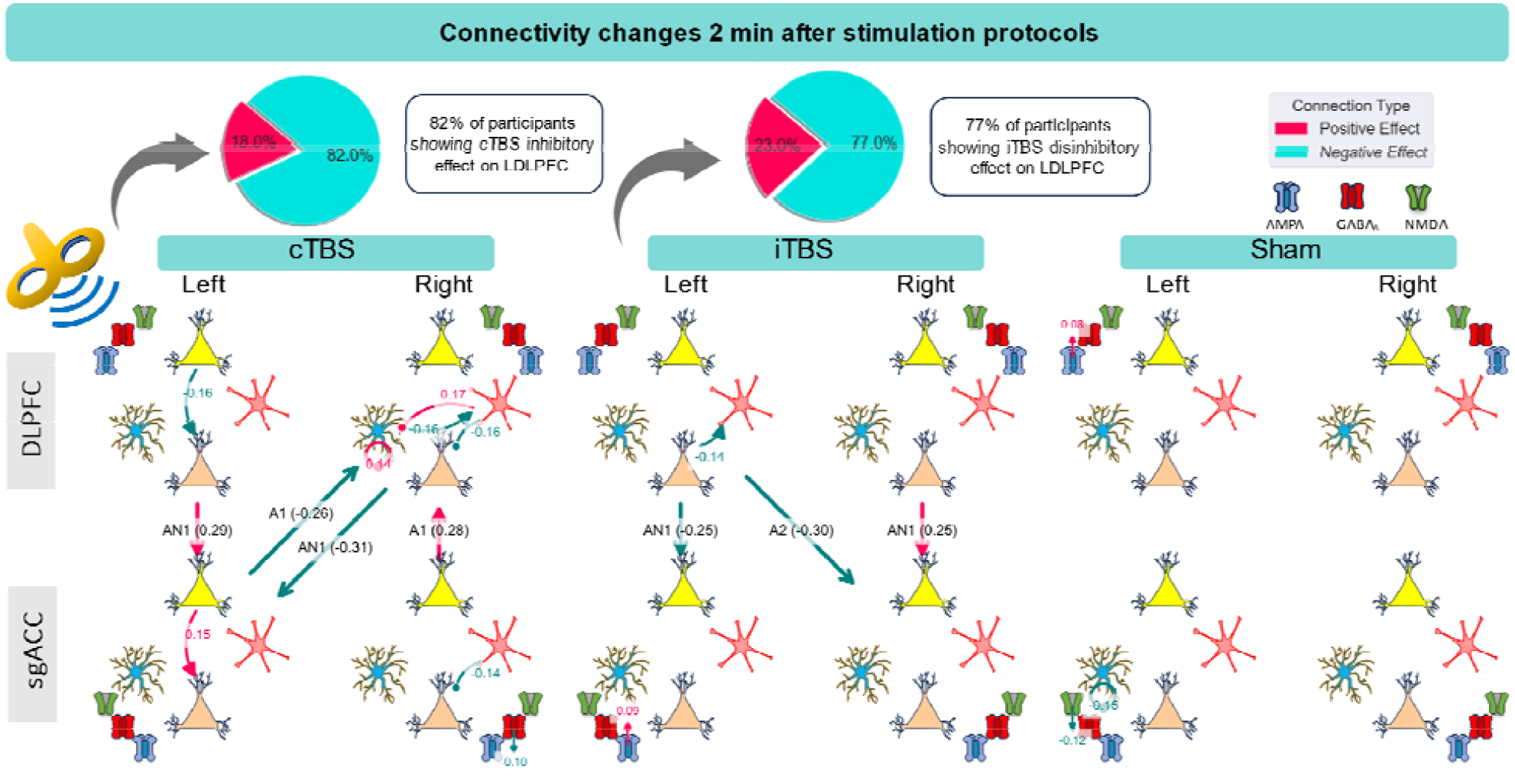
Connectivity changes two minutes after cTBS, iTBS, and sham (pre vs post1). Parameters with 95% or higher probability, indicating strong evidence, displayed for each protocol. Red and blue indicate positive (increasing connectivity) and negative (decreasing connectivity) effects. Receptor time constant (T) illustrate beside each source, where blue corresponds to AMPA, red to GABA_A_, and green to NMDA. A1: forward AMPA extrinsic connection, A2: backward AMPA extrinsic connection, AN1: forward NMDA extrinsic connection, AN2: backward NMDA extrinsic connection.

### Neural parameters that best explain TBS effect over time

To identify which neural parameters best explain TBS-induced changes across time, we combined each subject’s DCM estimates from all sessions into a single PEB and compared a series of reduced models. Each reduced model varied the inclusion of intrinsic properties and extrinsic connections, spanning Pre, Post1 (2 min), Post2 (15 min), and Post3 (30 min).

Bayesian model comparison revealed the full model, which simultaneously incorporates intrinsic microcircuit parameters, intrinsic self-connections, and extrinsic interregional coupling, as having the highest PEB free energy. This finding suggests that TBS induces hierarchical changes from interregional connectivity down to cellular dynamics (**Figure 5**).

**Figure 5.**
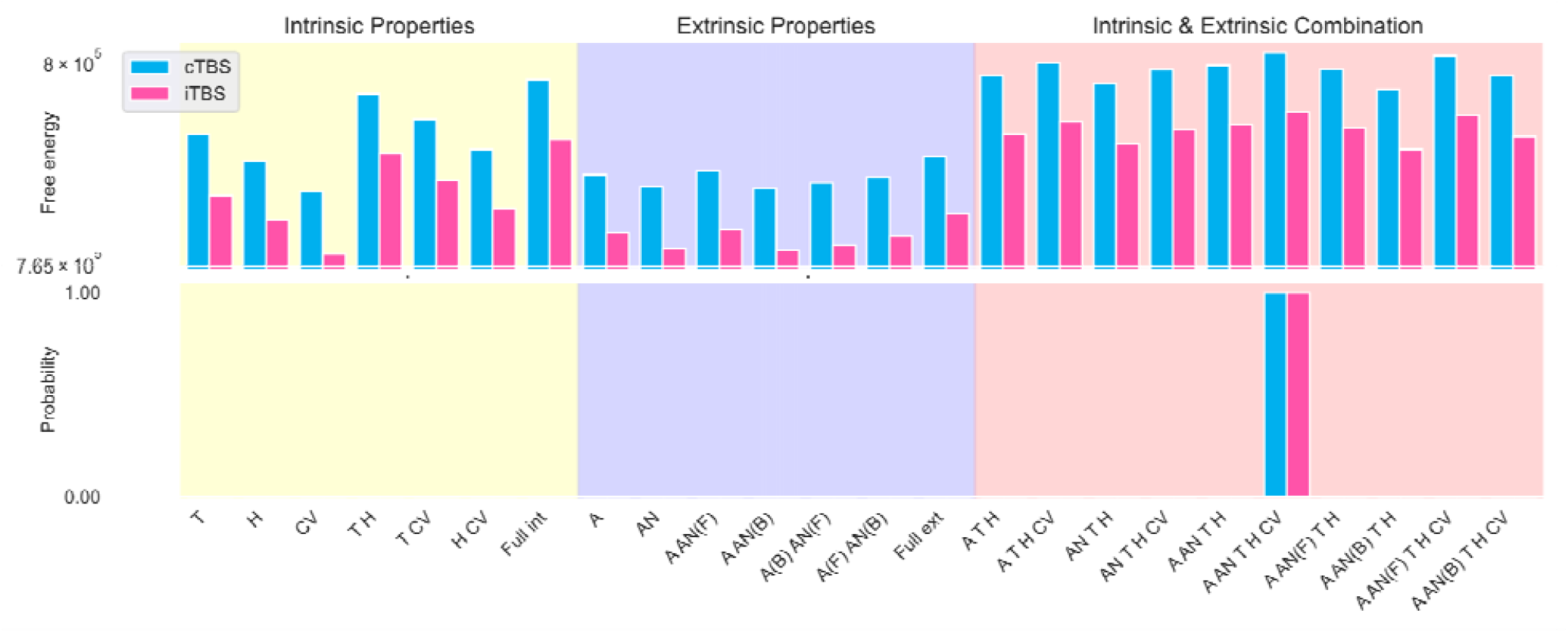
Bayesian model comparison showing the full model could best explain TBS differences. Bar plots show PEB free energy (top) and posterior model probability (bottom) for three families of reduced models—intrinsic properties only, extrinsic connections only, and combined of both. The full model, which integrates all the intrinsic and extrinsic features, achieved the highest evidence for both protocols, indicating that TBS effect could best explain with hierarchical changes from synaptic to network scales.

Based on prior rTMS literature demonstrating both sustained and transient neural effects^1,6,30^, we specified two temporal regressors: a sustained effect regressor ([−1 1 1 1]) to capture changes persisting up to 30 minutes post-stimulation, and a transient effect regressor ([−1 1 1 −1]) to detect changes that return to baseline after 30 minutes. Separate estimations for each protocol (iTBS, cTBS, sham) showed evidence for both sustained and transient modulation across protocols. To isolate parameters specific to each TBS condition, we performed between-protocol contrasts against sham using a design matrix of [−1 1]. This analysis highlighted protocol-specific intrinsic and extrinsic parameter changes, distinguishing neuromodulatory profiles of iTBS and cTBS beyond shared effects observed in sham (**Figure 3.B**).

### Sustained and transient changes following cTBS vs Sham

**Figure 6A** depicts the sustained and transient cTBS-induced changes relative to sham. We observed a transient reduction in NMDA and AMPA-mediated forward connectivity from right DLPFC to left sgACC, and in AMPA-mediated forward coupling from left sgACC to right DLPFC. Interestingly, iTBS produced similar transient decreases, indicating a common short-lived modulation for both TBS protocols beyond sham **(Table 2)**.

**Table 1.**
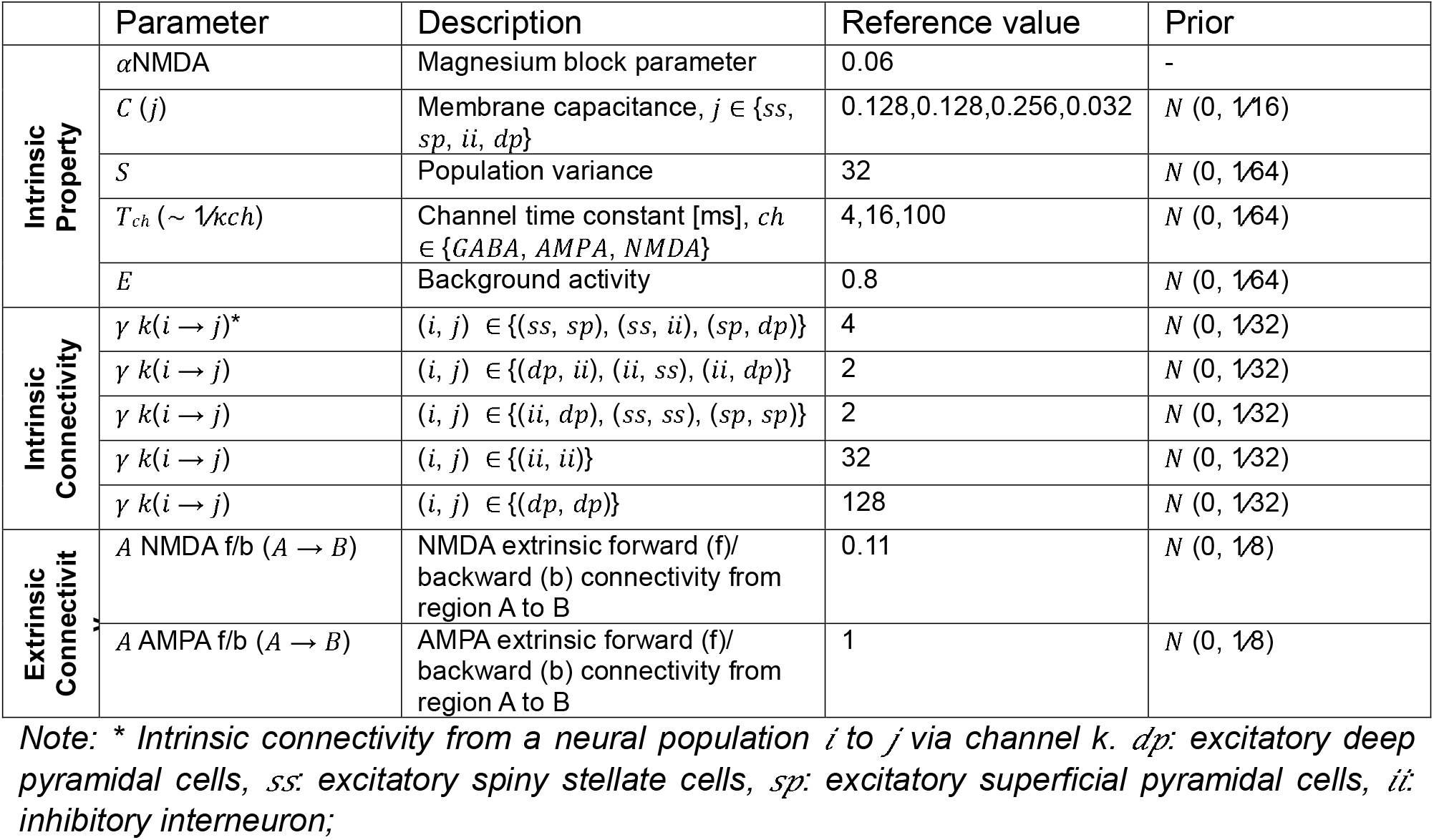
Description of parameters.

**Table 2.** PEB of PEB results; *Sustain and Transient changes following iTBS compared to Sham*.

| iTBS>Sham |  |  |  |
| --- | --- | --- | --- |
| Sustain |  | Transient |  |
| Parameters | $B_1$ | Parameters | $B_1$ |
| $T_{AMPA}(L.DLPFC)$ | -0.0944 | $T_{AMPA}(L.sgACC)$ | -0.0535 |
| $T_{AMPA}(L.sgACC)$ | 0.0952 | $A_{fAMPA}(L.DLPFC \rightarrow R.DLPFC)$ | 0.1860 |
| $T_{NMDA}(L.DLPFC)$ | 0.0644 | $A_{fAMPA}(L.sgACC \rightarrow R.DLPFC)$ | -0.1960 |
| $A_{bAMPA}(R.sgACC \rightarrow L.sgACC)$ | 0.1929 | $A_{fAMPA}(LsgACC \rightarrow R.sgACC)$ | 0.1838 |
| $H_{L.DLPFC}(II \rightarrow SP)$ | 0.0882 | $A_{bAMPA}(R.sgACC \rightarrow LsgACC)$ | -0.1945 |
| $H_{R.DLPFC}(SS \rightarrow SP)$ | 0.1517 | $A_{fNMDA}(R.DLPFC \rightarrow LsgACC)$ | -0.1647 |
| $H_{R.sgAAC}(SS \rightarrow II)$ | -0.0914 | $H_{R.DLPFC}(SS \rightarrow SP)$ | -0.1039 |
| $H_{R.sgAAC}(SP \rightarrow DP)$ | 0.0908 | $H_{L.sgAAC}(SS \rightarrow II)$ | -0.0830 |
| $H_{LsgACC}(DP \rightarrow II)$ | 0.1036 | $H_{LsgAAC}(II \rightarrow SS)$ | -0.0840 |
| $H_{L.sgAAC}(SS \rightarrow SS)$ | 0.100 | $H_{R.sgAAC}(SP \rightarrow DP)$ | -0.1032 |
| | | $H_{L.DLPFC}(SS \rightarrow II)$ | -0.0822 |

**Figure 6.**
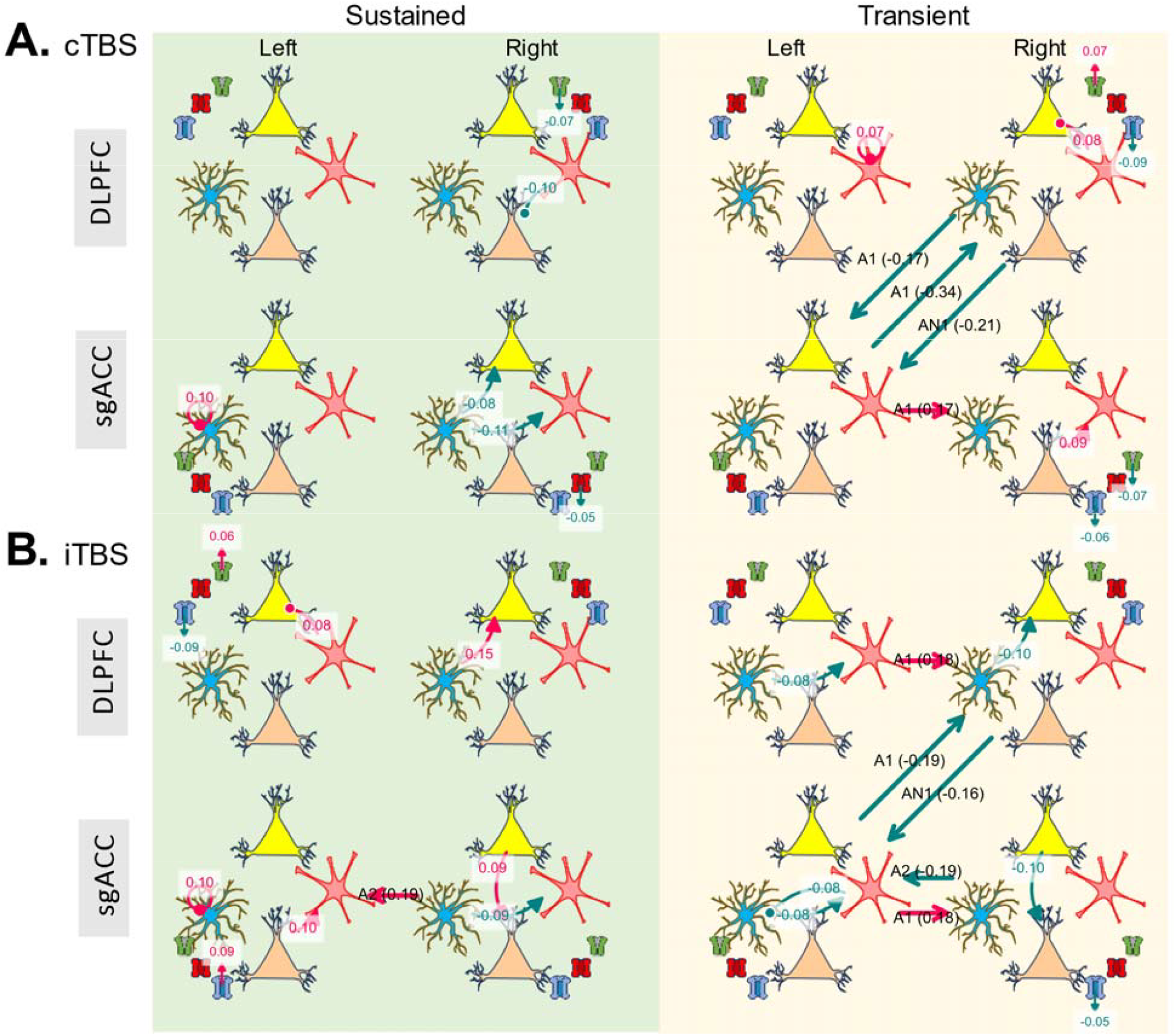
Sustain and transient changes following TBS compared to sham. (A) cTBS vs Sham (B) iTBS vs Sham. Sustained effects reflect lasting changes up to 30 min post-TBS, while transient effects capture the parameters altered post-TBS but returning to baseline by 30 min. Red and blue indicate positive (increasing connectivity) and negative (decreasing connectivity) effects. Receptor time constant (T) illustrate beside each source, where blue corresponds to AMPA, red to GABA_A_, and green to NMDA. A1: forward AMPA extrinsic connection, A2: backward AMPA extrinsic connection, AN1: forward NMDA extrinsic connection, AN2: backward NMDA extrinsic connection.

The inhibitory effect of cTBS in the left DLPFC was transient: we found increase II self-inhibition that returned to baseline by 30 minutes. No sustained intrinsic changes were observed in left DLPFC. However, right DLPFC exhibited a sustained reduction in II→DP, reflecting prolonged disinhibition. Extrinsic connectivity alterations following cTBS largely reverted to baseline by 30 minutes, with no enduring changes.

In the bilateral sgACC, microcircuit dynamics exhibited sustained hypoactivity after cTBS: left sgACC showed increased self-inhibition of SS cells, while right sgACC demonstrated decreased SS→SP and SS→II.

### Sustained and transient changes following iTBS vs Sham

**Figure 6B** illustrates iTBS-specific effects compared to sham. Most extrinsic connectivity modulations returned to baseline by 30 minutes, except for a sustained increase in AMPA-mediated backward connectivity from right sgACC to left sgACC. Intrinsic parameters revealed sustained changes in all four regions. In left DLPFC, II→SP increased, whereas right DLPFC showed increase excitatory connection from SS→SP. The left sgACC additionally showed local inhibition following iTBS, evidenced by increased self-inhibition in SS cells and enhanced excitatory connection from DP→II **(Table 3)**..

**Table 3.** PEB of PEB results; *Sustain and Transient changes following cTBS compared to Sham*.

| cTBS>Sham |  |  |  |
| --- | --- | --- | --- |
| *Sustain* |  | *Transient* |  |
| *Parameters* | *B<sub>1</sub>* | *Parameters* | *B<sub>1</sub>* |
| T<sub>GABA</sub>(R.sgACC) | -0.0558 | T<sub>AMPA</sub>(R.DLPFC) | -0.0913 |
| T<sub>NMDA</sub>(R.DLPFC) | -0.0723 | T<sub>AMPA</sub>(R.sgACC) | -0.0699 |
| H<sub>R.DLPFC</sub> (II → DP) | -0.1012 | T<sub>NMDA</sub>(R.DLPFC) | 0.0776 |
| H<sub>L.sgAAC</sub> (SS → SS) | 0.1004 | T<sub>NMDA</sub>(R.sgACC) | -0.0730 |
| H<sub>R.sgAAC</sub> (SS → SP) | -0.0842 | CV(L.DLPFC) | -0.1323 |
| H<sub>R.sgAAC</sub> (SS → II) | -0.1111 | CV(R.DLPFC) | 0.1151 |
|  |  | A<sub>f AMPA</sub> (R.DLPFC→LsgACC) | -0.1715 |
|  |  | A<sub>f AMPA</sub> (L.sgACC→R.DLPFC) | -0.3436 |
|  |  | A<sub>f AMPA</sub> (RsgACC → L.sgACC) | 0.1720 |
|  |  | A<sub>f NMDA</sub> (R.DLPFC→LsgACC) | -0.2200 |
|  |  | H<sub>L.DLPFC</sub> (II → II) | 0.0799 |
|  |  | H<sub>R.DLPFC</sub> (II → SP) | 0.0867 |
|  |  | H<sub>R.sgAAC</sub> (DP → II) | 0.0901 |

### Cellular property changes following TBS

Our model’s conductance-based architecture also permits estimation of receptor time constants for AMPA, NMDA, and GABA(A) channels. Compared to sham, iTBS induced sustained increases in NMDA time constants and decreases in AMPA time constants in left DLPFC, alongside prolonged increases in AMPA time constants in left sgACC. Conversely, cTBS produced sustained decreases in NMDA time constants in right DLPFC and reductions in GABA(A) time constants in right sgACC. Although channel dynamics interact nonlinearly with intrinsic connectivity^24^, these lasting receptor time constant changes in sgACC align with the observed microcircuit hypoactivity—namely, increased AMPA time constants after iTBS and reduced GABA(A) time constants after cTBS, both reflecting enduring inhibitory effects over sgACC that is absent in sham.

## Discussion

Spectral DCM with a canonical microcircuit model incorporating NMDA has recently gained increasing attention in computational neuropharmacology for assessing drug effects across hierarchical biological levels^24,31,32^. In this study, for the first time, we aimed to investigate TBS-induced changes in the DLPFC-sgACC microcircuit network using this approach in healthy adults. These regions have been repeatedly implicated in the pathophysiology of depression and are known to play a crucial role in mediating the antidepressant effects of rTMS^11,16,33–35^.

Recent TMS/fMRI studies consistently report that DLPFC stimulation elicits both acute and lasting changes in sgACC activity^14,15,36^. Trapp et al. provided the first direct electrophysiological evidence showing that sgACC activity is modulated by DLPFC stimulation, using TMS combined with intracranial EEG^16^. Given the role of sgACC hyperactivity in depression, the therapeutic effects of rTMS are thought to involve indirect regulatory modulation of this region ^18^. Furthermore, fMRI studies have reported that sgACC–DLPFC connectivity is enhanced in MDD patients^33,34^ and that successful rTMS treatment downregulates this connectivity^37^. Notably, we found that both TBS protocols, but not Sham, reduced right DLPFC to left sgACC effective connectivity that is NMDA-dependent. Although these extrinsic connectivity changes were transient, returning to baseline within 30 minutes post-stimulation, they induced lasting changes within the sgACC microcircuit.

Unlike Sham, both TBS protocols reduced left sgACC activity by increasing self-inhibition on spiny satellite cells and modulating receptor dynamics. Specifically, we found a decrease in the GABA-A time constant in the right sgACC following cTBS and an increase in the AMPA time constant in the left sgACC following iTBS. This suggests that dynamic changes in GABA-A and AMPA receptors also contribute to sgACC inhibition. We ensured that these changes were not driven by fatigue or other confounding factors by identifying sustained and transient changes that differed from Sham in the between-subject PEB design. This finding aligns with a large randomized clinical trial that analyzed source-localized TMS-evoked potentials in MDD patients following rTMS treatment over the left DLPFC^37^. In particular, they found that higher subgenual cingulate cortex (SGC) source activation and DLPFC-SGC connectivity, as measured by significant current scattering, were attenuated after active rTMS and significantly correlated with symptom improvement in MDD patients. Furthermore, they argued that this reduction in connectivity is mediated by GABAergic neurotransmission, based on the observation of an evoked potential at 100 milliseconds post-TMS pulse, associated with neuronal inhibitory processes^37^. Our results, using source localization of resting-state EEG data and a model-based effective connectivity approach, similarly found this effect in healthy individuals.

Additionally, we found evidence for an immediate inhibitory effect of cTBS and a disinhibitory effect of iTBS on the left DLPFC at the group level. However, within-subject analysis showed that these changes were not present in all participants, with some individuals even showing opposite effects. Initially, we hypothesized that these changes would persist for up to 30 minutes after stimulation. However, we found that these effects were transient, and the DLPFC microcircuit connections that changed and lasted over time did not follow this trend. In particular, iTBS induce lasting inhibition over stimulation target with increasing inhibitory connection to superficial pyramidal cells, while cTBS induce no sustain change over left DLPFC that differed from Sham. This finding speaks against the notion of a sustained inhibitory or excitatory effect of cTBS and iTBS over prefrontal regions. The local effects of TBS over the DLPFC stimulation target have also been mixed in neuroimaging studies^4,19^. For example, Shang et al. found no significant cerebral blood flow changes in the DLPFC following cTBS^38^, whereas Chang et al.^20^ reported an increased BOLD response in the stimulation target following iTBS. In a comprehensive review of concurrent TMS-fMRI studies, the authors argued that TMS does not consistently increase BOLD activity at the site of stimulation, perhaps due to alternating periods of increased and decreased neuronal firing that cancel each other out^39^. At the individual neuronal level, Romero et al.^6^ found non-uniform responses following cTBS over parietal neurons in awake, behaving rhesus monkeys. While they observed an overall inhibitory effect at the population level, peaking 30–40 minutes post-stimulation, transient phases of hyperexcitability were also noted^6^. Together, our findings challenge the assumed sustained effect of TBS protocols at the stimulation target and further demonstrate that the TBS effect is variable between subjects and dynamically changes over time.

Direct comparisons between cTBS and iTBS effects over time revealed both shared and distinct patterns. cTBS induced broader network-level effects at 2 minutes post-stimulation, consistent with previous neuroimaging findings^2^. In contrast, iTBS produced more sustained changes in intrinsic connectivity. Both protocols increased inhibition within the left sgACC and enhanced excitation in the right DLPFC; however, only iTBS led to lasting modulation at the stimulation target. A recent computational modeling study of longitudinal TMS-EEG data in 90 MDD patients undergoing iTBS treatment showed that iTBS suppresses low-frequency TMS-evoked potentials (TEPs) oscillatory power by modulating inhibition, with treatment responders exhibiting stronger inhibitory interneuron-to-pyramidal cell connections using the Jansen-Rit neural mass model^40^. In line with this, our results also revealed increased inhibition from inhibitory interneurons to superficial pyramidal cell (II→SP) in the left DLPFC following iTBS, but not after sham or cTBS. Furthermore, a meta-analysis of 23 randomized controlled trials comparing TBS protocols in depression found that iTBS over the left DLPFC was associated with greater symptom improvement and higher remission rates than cTBS. Together, these findings suggest that the superior clinical efficacy of iTBS may be driven by its ability to induce more durable intrinsic microcircuit changes, rather than simply acting as an excitatory protocol.

A recent DCM study by Pantazatos et al.,^41^ which modeled fMRI time-varying connectivity between the DLPFC and sgACC following TMS, found that TMS modulates both intrinsic connectivity as well as bottom-up and top-down connectivity between these regions. Aligning with this study, we found that our full model, incorporating a combination of intrinsic and extrinsic connections, could best explain the TBS effect. This clearly highlights the complex hierarchical neurobiological effects of TMS, creating a “chain of causation.”^22^ We further found that these neuromodulatory effects dynamically change over time. For example, a few parameters showed a negative change in the transient phase, followed by a positive shift in the sustained regressor (see **Figure 6**), suggesting a potential compensatory neural mechanism emerging during the later post-stimulation period (Post 3). A promising direction for future work would be to apply data-driven clustering methods to the estimated neural parameters to better capture possible connectivity changes over time.

This cascade of effects may be confounded by multiple factors, and we argue that such neural changes should not be attributed solely to TBS without high-temporal-resolution measurements, carefully controlled sessions, and robust statistical inference to capture dynamic processes over time. In our modeling approach, we could clearly observe these confounding factors reflected in the neural parameters. For example, previous studies have suggested that sgACC-induced activity following DLPFC stimulation might be explained by emotional (e.g., pain) or somatosensory experiences, due to the dense sgACC connections to limbic regions, including the amygdala and its role in affective processing^16^. Yet, the question of whether sgACC-induced activity is confounded by emotional or somatosensory experiences, or solely driven by DLPFC stimulation, cannot be easily captured by the slower and indirect nature of fMRI BOLD signals. Here, we found evidence for both. Immediately after stimulation, the only parameter change following the Sham protocol was observed in the left sgACC, where we found reduced self-inhibition in spiny satellite cells, indicating sgACC disinhibition. In the cTBS session, we also found bilateral sgACC hyperactivity after Post1. We believe these early changes, particularly following Sham and cTBS, could be better interpreted as an initial emotional or sensory reaction rather than direct neuromodulatory effects of stimulation. However, including parameters from post 2 (−15 minutes after stimulation) and post 3 (−30 minutes after stimulation) in our PEB analysis, and comparing them with Sham, revealed sustained sgACC inhibition following TBS that was not present in Sham. This finding could support the previously proposed antidepressant effect of TBS on reducing sgACC hyperactivity^10,17,37^. Future studies are necessary to replicate these results in MDD patients and, more importantly, to evaluate how these parameters could predict clinical improvements.

The current dataset also includes TEPs recorded following the TBS protocols. Analysis of TEP component amplitudes within a predefined left DLPFC ROI (i.e., average of F3, FC1, FC3) has been reported elsewhere, showing no significant differences between active and sham conditions^42^. Our present study, by contrast, focuses on resting-state EEG using a fundamentally different analytic pipeline. Although TEP components have been proposed as sensitive markers of cortical excitability, their underlying electrophysiological mechanisms remain poorly understood, and they exhibit substantial inter-individual variability^4^. In contrast, the biological plausibility of spDCM and neural mass models offers a more nuanced and mechanistic account of cortical excitability at the microcircuit level^26^. Additionally, spDCM approach using the CMM-NMDA model has recently demonstrated strong reliability and reproducibility in resting-state electrophysiology^28^, and the multilevel Bayesian approach (PEB of PEB) incorporates group-level informative priors at both the within-subject and between-subject levels to address heterogeneity across individuals^29^. While we argue that this technique is especially promising for tracking dynamic neurophysiological responses to TMS, several methodological challenges remain. First, the choice of window length in resting state EEG for spDCM analysis has not been well established^43^; we used a 2-second window here following^24^, although prior studies have also applied 1-second^28,43^ 5-second^31^ or even 10-second^44^ window. Additionally, the current neuronal model cannot explain how on-level changes affect other neural parameters (e.g., how changes in time constants affect intrinsic connectivity, and vice versa), making the model blind to temporal orders^24^. Addressing this issue would require a biologically grounded model for the trans-hierarchical progression of TBS effects rather than the current all-in-one CMM-NMDA model^24^. Nonetheless, given the non-invasive nature, low cost, and portability of resting-state EEG compared to TMS-fMRI or TMS evoked potentials, this method holds considerable promise for tracking time-dependent changes in brain connectivity in future neuromodulation research.

There are important limitations to consider when interpreting the results. First, our finding only examines DLPFC-sgACC connectivity in healthy individuals, which may not be replicable in MDD patients. Although previous studies have shown that DLPFC-sgACC functional connectivity from normative connectomes can predict clinical response comparably to disease-matched connectomes^10,11^, differences in connectivity have been reported in patients with depression^33,34,37,41^. For example, Pantazatos et al. showed that TMS modulates DLPFC-sgACC effective connectivity differently in healthy individuals compared to patients^41^. Future studies are needed to directly compare the current results in clinical populations. Second, TBS was applied at 75% of the RMT, following Chung et al.^45^, who reported the strongest iTBS effects in the prefrontal cortex at this intensity. However, it is important to note that this relatively low stimulation intensity may limit comparability with clinical protocols, which typically use 100% or higher.

With these limitations in mind, we have uncovered protocol-specific and receptor-mediated dynamics of TBS between the DLPFC and sgACC by using spectral DCM with biophysically realistic microcircuit models. These results provide mechanistic insight into how TBS may achieve therapeutic modulation of the sgACC and offer a powerful computational toolkit for future precision neuromodulation studies.

### Method Dataset

The EEG recordings analyzed in this study were obtained from a peer-reviewed open-source dataset^46^ (Available: https://doi.org/10.25452/figshare.plus.c.5910329). This dataset employed a single-blinded, sham-controlled crossover design. Twenty four healthy right-handed participants (11 females with mean age of 25.2 ± 9.9 years) attended five sessions scheduled at the same time of day, each separated by at least one week to minimize carryover effects. Session order was pseudo-randomized via computer-generated sequences: three initial sessions, each comprising one of iTBS, cTBS, or sham (randomized), followed by two additional sessions (one each of iTBS and cTBS). Participants were excluded if they met any of the following criteria: (1) a history of psychiatric or neurological disorders, including seizures or stroke; (2) recent head injury; (3) current use of medications that affect cognitive function; (4) drug or alcohol abuse within the past three months; (5) smoking; (6) pregnancy; or (7) any contraindications to EEG or MRI.

Resting motor thresholds (RMTs) were determined individually by identifying the minimum stimulus intensity required to evoke at least 3 out of 6 motor-evoked potentials (MEPs) with a peak-to-peak amplitude of ≥50□µV in the contralateral right first dorsal interosseous muscle. Electromyographic (EMG) recordings were obtained using a 1401 laboratory interface and 1902 amplifier (Cambridge Electronic Design, Cambridge, UK), with data acquired via the Signal V4 software (Cambridge Electronic Design).

TBS and single-pulse TMS were administered using a MagPro® X100 stimulator (MagVenture, Lucernemarken, Denmark) equipped with a 65 mm Cool-B65 figure-8 coil. The coil was positioned tangentially to the scalp over the F3 electrode location to target the left DLPFC, and TBS was delivered at 75% of each individual’s RMT^46^. For the iTBS protocol, stimulation consisted of two-second trains delivered every 10 seconds over 192 seconds, totaling 600 pulses^5^. The cTBS protocol involved 600 uninterrupted pulses delivered over 40 seconds^5^. The sham condition used an inactive coil placed directly on the scalp, while a second active coil, positioned 20 cm posterior and oriented away from the head, was used to simulate the TMS clicking sound. This posterior coil operated at 20% higher intensity to account for the increased distance from the ear and delivered either an iTBS or cTBS pattern (randomly assigned across participants). To further enhance blinding, participants wore earphones playing white noise at a volume sufficient to mask the auditory artifact of the stimulation, adjusted to the highest level tolerable for each individual^46^.

Four minutes of eyes-open resting-state EEG were recorded at the session’s start and after completing all four blocks of single-pulse TMS, as illustrated in **Figure 2**. EEG data were recorded using a Refa amplifier (2048 Hz sampling rate; TMSi, Oldenzaal, The Netherlands) and a 64-channel 10–20 EEG cap (EasyCap, GmbH, Herrsching, Germany), with cap size chosen per participant. Electrodes were grounded at Fpz and captured with a common average reference. Recordings took place in a dimly lit, sound-attenuated room, with participants fixating on a cross displayed on a 58.4 cm (23-inch) monitor.

### EEG preprocessing

The data were processed and analyzed offline. This was done using a combination of open-source toolboxes, including Fieldtrip^47^, EEGLAB (version 2022b)^48^, and custom scripts, all implemented on the MATLAB platform (R2022b, The MathWorks, USA).

Eyes-open resting-state EEG data were preprocessed by down-sampling to 256 Hz, baseline correction (demeaning), and detrending to remove any linear trends. A second-order bandpass filter (1–50 Hz) and a 50 Hz notch filter were applied to eliminate electrical line noise. The data were segmented into 2-second epochs, with an automated algorithm first rejecting epochs exhibiting data ranges exceeding three standard deviations or absolute maxima surpassing 12 standard deviations of other epochs. Remaining noisy epochs were excluded through visual inspection. Independent Component Analysis (ICA) was performed once to identify and remove components associated with eye blinks, eye movement and muscle artifacts. The data were then re-referenced to the common average reference. One of the subjects exclude at this stage due to poor quality in Sham session.

### Dynamic causal modelling for resting state EEG

Preprocessed data were imported into SPM25^49^ (revision 25.01.rc3) to perform DCM for cross-spectral densities (CSD) (https://www.fil.ion.ucl.ac.uk/spm/). This steady-state variant uses a conductance-based canonical microcircuit model (cmm_NMDA) to estimate how endogenous neuronal fluctuations produce the observed CSD^27^. DCM inverts a generative neural mass model alongside an observation model to infer intrinsic and extrinsic connectivity parameters that best explain the empirical data^27^.

### Conductance-Based Neural Mass Model

The Conductance-Based Neural Mass Model with NMDA (CMM-NMDA) is a variant of electrophysiological DCM. This model describes the dynamic behavior of neural populations by incorporating the conductance of three primary ionotropic receptors: AMPA, GABA-A, and NMDA receptors. The conductance of these channels is determined by the cumulative synaptic interactions within and between cortical regions. In the CMM-NMDA framework, neural activity emerges from four key neural populations: excitatory spiny stellate cells (SS), superficial pyramidal cells (SP), inhibitory interneurons (II), and deep pyramidal cells (DP). These populations are interconnected via intrinsic connections within the cortical regions. Moreover, the model captures the extrinsic connectivity between cortical regions^27^. We can describe the CMM-NMDA model in 3 levels: intrinsic properties, intrinsic connections, and extrinsic connections (**Figure 1**).

### Neural population properties

Each population in a cortical region is modelled by a Morris–Lecar model^50^ and contains four hidden states: the membrane voltage *V* and the conductance *gk* for all three receptors: AMPA, GABA-A, and NMDA^51,52^. The mean membrane voltage of population j is obtained by this equation:

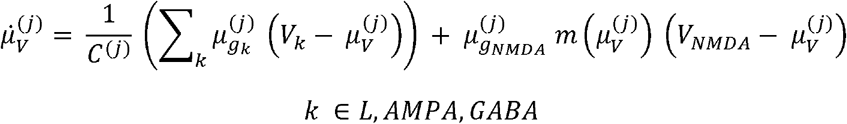

Where:

*j* is the target neural population;

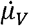 is the time derivative of mean voltage;

*μ*_*V*_ is the mean voltage;

*C* is the membrane capacitance;

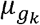 is the mean conductance of channel k;

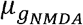 is the mean conductance of the NMDA channel;

*V*_*k*_ is the reversal potential of channel k;

*V*_*NMDA*_ is the mean reversal potential of the NMDA channel;

L is a passive leak channel, which means its conductance is a fixed number (1 in SPM25). Each channel’s conductance is voltage-time-dependent. The mean conductance of channel k of population j is obtained by this equation:

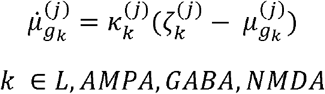

Where:

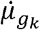 is the time derivative of the mean conductance of channel k;

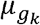 is the mean conductance of channel k;

κ_*k*_ is the inverse time constant of channel k;

ζ_*k*_ is the input to the target neural population;

NMDA has a voltage-dependent magnesium block, too:

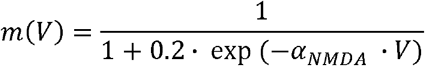

Where *α*_*NMDA*_ is the magnesium block parameter, which is a fixed parameter (Table 1).

### Intrinsic connection

The populations are interconnected via intrinsic connections within the cortical regions. The input to the target population is the cumulative output of activated origin populations:

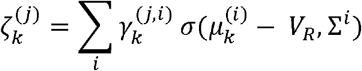

Where:

*j* is the target neural population;

*i* is the origin neural population;

*ζ_k_* is the input to the target neural population via channel k;

*γ_k_* is the intrinsic coupling parameter from origin to target neural population via channel k;

*α* is the function for proportion of activated neurons in the origin population and is formulated as a cumulative distribution of the univariate normal distribution 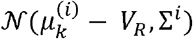 where *V*_*R*_ is the threshold potential (−40 in SPM25) and ∑^*i*^ is a free parameter.

### Extrinsic connection

Extrinsic connections are divided into 2 types: Forward connections, which run from superficial pyramidal cells of lower cortical regions to spiny stellate cells of higher regions. Backward connections, which run from deep pyramidal cells of higher to superficial pyramidal and inhibitory interneurons of lower brain regions.

### Model specification and inversion

Anatomical coordinates of network nodes were defined a priori. Individual T1-weighted MRI scans were utilized to construct subject-specific head models and lead fields, ensuring accurate source localization. We specified a fully connected network comprising bilateral DLPFC and sgACC, modeling each region as a cortical patch using the “IMG” option in SPM25. Nodes were placed at the following MNI coordinates: left DLPFC^1^0 (−38, 44, 26), right DLPFC (38, 44, 26), left sgACC (−10, 20, −15) and right sgACC (5, 15, −15)^53^. For each subject, the individuals high-resolution T1-weighted MRI was first reoriented and co-registered to EEG electrode coordinates of that session using fiducial landmarks (nasion and preauricular points) and head-shape points, ensuring precise alignment of sensor and anatomical space. The MRI was then segmented into scalp, skull and brain compartments to construct an individual three-layer Boundary Element Method (BEM) head model, and a fine-resolution cortical mesh (mesh size = 3) was generated for accurate forward modeling and source localization^54^. Before inversion, data registration and mesh quality were visually inspected and saved for each session.

DCM Model inversion was performed using variational Bayes under the Laplace approximation, allowing for the estimation of parameters governing both neural dynamics and the observation model^51,55^. Parameter estimation involved maximizing the variational free energy defined as:

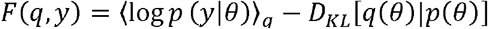

where *p* (*y* ∣ θ) represents the likelihood function, and *D*_*KL*_ is the Kullback–Leibler divergence between the recognition density *q* (*θ*) and the prior distribution *p* (*θ*) recognition density *q* (*θ*) provides an approximate posterior distribution Model fit was *p* (*y* ∣ θ) assessed by explained variance; ; Which compute as:

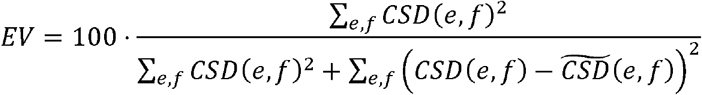

Where *CSD*(*e,f*) denotes the observed cross-spectral density (CSD) at frequency f for edge e, and 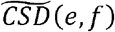 is the model-estimated CSD at the same frequency and edge. Participants with explained variance below 50% were excluded, resulting in removal of one subject prior to group-level analysis **(Supplementary Material, Figure S1)**.

### Group level analysis using multilevel parametric empirical bayes

Following DCM parameter estimation, we applied a parametric empirical Bayesian (PEB) framework with SPM25 to examine systematic variations in model parameters induced by TBS and Sham over 22 subjects.

While an alternative approach would involve applying classical statistics (e.g., repeated-measures ANOVA) to the expected posterior means (Ep), this method overlooks the underlying uncertainty associated with DCM parameters^56^. Since DCM estimates are multivariate normal distributions, relying solely on point estimates disregards the posterior covariance (Cp), which reflects the confidence in parameter estimation^56^. In contrast, the PEB framework incorporates both Ep and Cp in group-level inference. This means that participants with more precise parameter estimates contribute more strongly to the group result, whereas those with noisier data are down-weighted^56^. This property is particularly advantageous in clinical or high-variability datasets, where reliable individual-level inference is critical.

A further challenge in working with nonlinear and ill-posed models—such as neural mass models applied to resting-state EEG—is the risk of converging on different local optima when fitting multiple full DCMs across sessions and subjects^29^. This issue emphasizes the importance of hierarchical modeling frameworks like PEB that can stabilize inferences despite local variations in model fits. To address the potential for session-level variability and local optima in DCM inversion, we used a multilevel PEB framework specifically adapted for longitudinal DCM analysis^29^. In this hierarchical model, within-subject longitudinal effects across sessions were modeled at the second level, and between-subject effects were modeled at the third level.

Empirical priors derived at each level were used to iteratively optimize posterior densities over parameters in the level below. At the first level, subject- and session-specific spDCMs were estimated. At the second and third levels, Bayesian general linear models were used to capture within- and between-subject effects, respectively^29^. The model hierarchy can be expressed as:

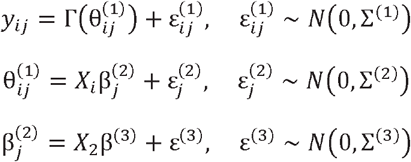

In the first equation, the observed time series data *yij* for the *ith* session and *jth* subject are modeled as the output of a nonlinear DCM function Γ plus observation noise 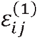.The model parameters 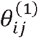 are sampled from the subject-specific second-level model *jth*, which incorporates session-level effects (pre, post1, post2, post3) with design matrix *X*1 and parameters 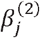, with added noise 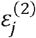.

The within-subject design matrix *X*_1_ used in our study was defined as: *X*_1_= [1 1 1 1; −1 1 1 1; −1 1 1 −1]. This allowed us to separate baseline, sustained, and transient post-TBS effects for each protocol. At the third level, subject-specific effects 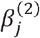 were modeled using the between-subject design matrix *X*_2_ and group-level parameters 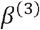.To compare TBS with Sham conditions across all subjects, we used: *X*_2_= [1 1; −1 1]. This design tested for common and differential effects of TBS relative to Sham (see **Figure 3**).

In this approach, extrinsic, intrinsic, and cellular parameters 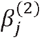 can be modeled as either fixed or random effects by specifying appropriate priors on between-subject variability in the hierarchical model^56^. To determine which parameters should be treated as random effects, we performed Bayesian model comparison by evaluating the free energy of reduced models using the “softmax” function^25^. This approach offers two key advantages: (1) it quantifies the contribution of each neural parameter in explaining TBS effects, and (2) it identifies the simplest model that best explains group differences.

We explored three classes of hypotheses regarding the parameters: (1) TBS effects are best explained through changes in extrinsic (between-region) connections; (2) TBS effects are better accounted for by modulations in intrinsic (within-region) microcircuit parameters; and (3) TBS affects both intrinsic and extrinsic connectivity. The model yielding the highest free energy was selected and treated as a random effect (“field” in the spm_dcm_peb function) for subsequent analyses.

Following the estimation of group-level effects, we employed automatic model search using Bayesian model reduction (BMR) and Bayesian model averaging (BMA) via the spm_dcm_peb_bmc function^56^. This greedy search algorithm iteratively removes parameters that do not contribute to model evidence. BMA was then performed across the 256 reduced models from the final iteration. Comparing full and reduced models in this way is conceptually similar to conducting a post-hoc F-test in classical statistics and is referred to as ‘post-hoc’ DCM analysis^56^. Lastly, it is important to note that group-level Bayesian analysis does not rely on the concept of significance in classical statistics. Instead, it provides posterior probabilities for each parameter. However, it is a common practice in DCM studies to report the most probable effects with posterior probability > 95, labelled as strong evidence based on Kass and Raftery^57^.

## Data and code availability

Raw EEG data available here: https://doi.org/10.25452/figshare.plus.c.5910329. Complete MATLAB and Python code used to estimate DCM and plotting parameters are available in the Github repository: https://github.com/ArminTi/Hierarchical-neurobiological-changes-in-DLPFC-sgACC-connectivity-induced-by-Theta-Burst-Stimulation/tree/main

## Authors contribution

A.T. conceptualized the study, analyzed the data, and wrote the manuscript. G.G. analyzed the data, created graphical illustrations, and assisted with manuscript review and editing. S.Z. participated in data analysis and manuscript editing. B.A. and H.M. contributed to data analysis.

A.N. supervised the project.

